## Supplementary Materials for "Psychosocial family-level mediators in the intergenerational transmission of trauma: Protocol for a systematic review and meta-analysis"

| Search term | Yield |
|---|---|
| ((intergenerational trauma.mp) or (transgenerational trauma.mp)) and ((systematic review.mp.) or (scoping review.mp)) | 27 |

**Table 1a**. Search strategy used to identify relevant systematic and scoping reviews in the Ovid (Medical Literature Analysis and Retrieval System Online (MEDLINE)® 1946 to February 03 2021, Embase 1974 to 2021 February 03, and APA PsycInfo 1806 to January Week 4 2021 databases. Date of search: 5 February 2021.

| Search term | Yield |
|---|---|
| Intergenerational trauma | 9 |

**Table 1b**. Search strategy used to identify reviews currently in progress in PROSPERO (https://www.crd.york.ac.uk/prospero) systematic review registration database. Date of search: 4 March 2021.

| Search term | Yield |
|---|---|
| “intergenerational trauma” | 2 |
| “transgenerational trauma” | 0 |

**Table 1c**. Search strategy used to identify reviews registered as an Open Science Framework preprint database (https://osf.io/preprints/). Date of search: 4 March 2021.

**eTable 1**. Search strategies used to identify review articles that highlighted psychosocial mediators or moderators in the intergenerational transmission of trauma. We also included papers from the first author’s personal library identified during initial screening (N=14).

| Reference | Review type | G1 and G2 population and trauma type restriction(s) | Generational scope | Number of quantitative studies that appear to measure psychosocial mediators and/or moderators on offspring mental health |
|---|---|---|---|---|
| *Peer-reviewed publications* | | | | |
| Cerdeña et al., 2021(1) | Scoping review | >5% Latinx populations in the United States and Canada who experienced "trauma or violence" | G1 and G2 | 15 (7 longitudinal; 1 interventional; 7 cross-sectional) |
| Dashorst et al., 2019(2) | Scoping review | Holocaust survivors | G1 and G2 | 23 (all quantitative studies; design unspecified) |
| Dekel et al., 2008(3) | Literature review | Male combat veterans | G1 and G2 | 6 (design unspecified) |
| Der Sarkissian & Sharkey, 2021(4) | Scoping review | Armenian genocide survivors | G1, G2, and G3 | Not reported |
| Flanagan et al., 2020(5) | Scoping review | Refugees with direct exposure to war-related trauma | G1 and G2 | 8 (2 longitudinal; 6 cross-sectional) |
| Galler & Rabinowitz, 2014(6) | Literature review | Parental early adversity and trauma | Unspecified; papers focused on G1 and G2 | 1 (cross-sectional) |
| Gone et al., 2019(7) | Scoping review | Historical trauma among indigenous populations | G1, G2, and G3 | 11 (design unspecified) |
| Kaitz et al., 2009(8) | Literature review | Parents traumatized by terror | G1 and G2 | Not reported |
| Kellerman, 2001(9) | Literature review | Holocaust survivors | Unspecified | Not reported |
| Leen-Felder et al., 2013(10) | Scoping review | Parental post-traumatic stress | G1 and G2 | 17 (design unspecified) |
| Lehrner & Yehuda, 2018(11) | Literature review | Holocaust survivors | G1, G2, and G3 | Not reported |
| Morris et al., 2012(12) | Systematic review and moderation meta-analysis | Parental and offspring post-traumatic stress symptoms | G1 and G2 | 32 (cross-sectional and longitudinal) |
| Pearrow & Cosgrove, 2009(13) | Literature review | Combat-related PTSD among military personnel | G1 and G2 | Not reported |

| | | | | |
|---|---|---|---|---|
| Plant et al., 2018(14) | Systematic review | Maternal childhood trauma | G1 and G2 | 12 (10 longitudinal; 2 cross-sectional) |
| Sangalang et al., 2017(15) | Scoping review | Refugees with trauma “collective in nature and affected a targeted group of people bound by a common cultural identity” | G1 and G2 | 16 (1 longitudinal; 15 cross-sectional) |
| Smallwood et al., 2021(16) | Scoping review | Historical trauma among indigenous populations | G1, G2, and G3 | 6 (3 longitudinal; 2 cross-sectional; 1 case-control) |
| Su et al., 2020(17) | Systematic review and moderation meta-analysis | Maternal childhood maltreatment | G1 and G2 | 12 (longitudinal) |
| Van Ee et al., 2016(18) | Scoping review | Caregivers with PTSD | G1 and G2 | 72 (design unspecified) |
| *Grey literature* | | | | |
| Kandlur 2021(19) | Rapid scoping review | Historical trauma among Native American communities | Unspecified | No quantitative studies identified |
| Constantino-Pettit, 2019(20) | PROSPERO registration for a systematic review; meta-analytic approach not reported | Mothers with childhood trauma | G1 and G2 | Unavailable |
| Khan & Rumesa, 2020(21) | Literature review, published as a conference abstract | Childhood trauma | G1 and G2 | Not reported |

**eTable 2**. Literature reviews of quantitative studies on potential psychosocial mediators and moderators for the intergenerational transmission of trauma.

| Section and topic | No | Checklist item | Location in Manuscript |
|---|---|---|---|
| ADMINISTRATIVE INFORMATION | | | |
| Title: | | | |
| Identification | 1a | Identify the report as a protocol of a systematic review | Title |
| Update | 1b | If the protocol is for an update of a previous systematic review, identify as such | Not applicable |
| Registration | 2 | If registered, provide the name of the registry (such as PROSPERO) and registration number | METHODS: paragraph two |
| Authors: | | | |
| Contact | 3a | Provide name, institutional affiliation, e-mail address of all protocol authors; provide physical mailing address of corresponding author | Title page |
| Contributions | 3b | Describe contributions of protocol authors and identify the guarantor of the review | AUTHORS' CONTRIBUTIONS |
| Amendments | 4 | If the protocol represents an amendment of a previously completed or published protocol, identify as such and list changes; otherwise, state plan for documenting important protocol amendments | METHODS: paragraph two |
| Support: | | | |
| Sources | 5a | Indicate sources of financial or other support for the review | FUNDING |
| Sponsor | 5b | Provide name for the review funder and/or sponsor | Not applicable |
| Role of sponsor or funder | 5c | Describe roles of funder(s), sponsor(s), and/or institution(s), if any, in developing the protocol | Not applicable |
| INTRODUCTION | | | |
| Rationale | 6 | Describe the rationale for the review in the context of what is already known | BACKGROUND, paragraphs 6 and 7 |
| Objectives | 7 | Provide an explicit statement of the question(s) the review will address with reference to participants, interventions, comparators, and outcomes (PICO) | REVIEW QUESTIONS |
| METHODS | | | |
| Eligibility criteria | 8 | Specify the study characteristics (such as PICO, study design, setting, time frame) and report characteristics (such as years considered, language, publication status) to be used as criteria for eligibility for the review | METHODS: Inclusion criteria |
| Information sources | 9 | Describe all intended information sources (such as electronic databases, contact with study authors, trial registers or other grey literature sources) with planned dates of coverage | METHODS: Protocol registration and timeline, Searches, and Source of evidence selection |
| Search strategy | 10 | Present draft of search strategy to be used for at least one electronic database, including planned limits, such that it could be repeated | Supplementary materials, eTables 3-8 |
| Study records: | | | |
| Data management | 11a | Describe the mechanism(s) that will be used to manage records and data throughout the review | METHODS: Source of evidence selection, Data extraction |

| | | | |
|---|---|---|---|
| Selection process | 11b | State the process that will be used for selecting studies (such as two independent reviewers) through each phase of the review (that is, screening, eligibility and inclusion in meta-analysis) | METHODS: Source of evidence selection, Data extraction |
| Data collection process | 11c | Describe planned method of extracting data from reports (such as piloting forms, done independently, in duplicate), any processes for obtaining and confirming data from investigators | METHODS: Data extraction |
| Data items | 12 | List and define all variables for which data will be sought (such as PICO items, funding sources), any pre-planned data assumptions and simplifications | METHODS: Data extraction and eTable 9 |
| Outcomes and prioritization | 13 | List and define all outcomes for which data will be sought, including prioritization of main and additional outcomes, with rationale | Not applicable, as mediators and moderators are the variables of interest |
| Risk of bias in individual studies | 14 | Describe anticipated methods for assessing risk of bias of individual studies, including whether this will be done at the outcome or study level, or both; state how this information will be used in data synthesis | METHODS: Meta-analyses: Assessment of methodological quality |
| Data synthesis | 15a | Describe criteria under which study data will be quantitatively synthesised | METHODS: Meta-analyses, paragraph 1 |
| | 15b | If data are appropriate for quantitative synthesis, describe planned summary measures, methods of handling data and methods of combining data from studies, including any planned exploration of consistency (such as $I^2$, Kendall's τ) | METHODS: Meta-analyses: Mediation meta-analyses, Moderation meta-analyses |
| | 15c | Describe any proposed additional analyses (such as sensitivity or subgroup analyses, meta-regression) | METHODS: Meta-analyses: Mediation meta-analyses, Moderation meta-analyses |
| | 15d | If quantitative synthesis is not appropriate, describe the type of summary planned | Not applicable |
| Meta-bias(es) | 16 | Specify any planned assessment of meta-bias(es) (such as publication bias across studies, selective reporting within studies) | METHODS: Meta-analyses: Publication bias |
| Confidence in cumulative evidence | 17 | Describe how the strength of the body of evidence will be assessed (such as GRADE) | METHODS: Assessing confidence in cumulative evidence |

**eTable 3**. PRISMA-P (Preferred Reporting Items for Systematic review and Meta-Analysis Protocols) 2015 checklist.(22)

| Search term | | Yield |
|---|---|---|
| 1 | (intergeneration* or transgeneration* or multigeneration* or crossgeneration* or generation* or secondary or secondarily or tertiary).mp.[intergenerational concept] | 1687825 |
| 2 | intergenerational relations/ or exp adult survivors of child adverse events/ or exp parent-child relations/ [intergenerational concept] | 63442 |
| 3 | (paternal* or maternal* or parent* or grandparent* or grandmother* or grandfather* or famil* or mother* or father* or child* or grandchild* or granddaughter* or grandson* or daughter* or son* or intergenerational* or transgenerational* or multigenerational* or generation* or caregiver* or mother-child).mp. [family setting] | 4699532 |
| 4 | ((intergeneration* or transgeneration* or multigeneration* or crossgeneration* or generation* or secondary or secondarily or tertiary or historical or vicarious or empathetic or legacy or inherited or maternal or paternal or parent* or mother* or father*) and (trauma* or PTSD or posttrauma* or psychotrauma* or abus* or maltreat* or mistreat* or adverse childhood or ACE or ACEs or childhood adversity)).ti. [trauma concept] | 6826 |
| 5 | ((intergeneration* or transgeneration* or multigeneration* or crossgeneration* or generation* or secondary or secondarily or tertiary or historical or vicarious or empathetic or legacy or inherited or maternal or paternal or parent* or mother* or father*) and (trauma* or PTSD or posttrauma* or psychotrauma* or abus* or maltreat* or mistreat* or adverse childhood or ACE or ACEs or childhood adversity)).kf. [trauma concept] | 2346 |
| 6 | ((intergeneration* or transgeneration* or multigeneration* or crossgeneration* or generation* or secondary or secondarily or tertiary or historical or vicarious or empathetic or legacy or inherited or maternal or paternal or parent* or mother* or father*) adj4 (trauma* or PTSD or posttrauma* or psychotrauma* or abus* or maltreat* or mistreat* or adverse childhood or ACE or ACEs or childhood adversity)).mp. [trauma concept] | 17131 |
| 7 | (1 or 2) and 3 and (4 or 5 or 6) | 5858 |

Date of search: January 21 2021.

**eTable 4**. Final search strategy to identify epidemiological studies assessing the intergenerational transmission of trauma at the family- and parent-level in the Ovid Medical Literature Analysis and Retrieval System Online (MEDLINE)® 1946 to Present database.

| Search term | | Yield |
|---|---|---|
| 1 | (intergeneration* or transgeneration* or multigeneration* or crossgeneration* or generation* or secondary or secondarily or tertiary).mp. | 206504 |
| 2 | intergenerational relations/ or transgenerational patterns/ | 7549 |
| 3 | (paternal* or maternal* or parent* or grandparent* or grandmother* or grandfather* or famil* or mother* or father* or child* or grandchild* or granddaughter* or grandson* or daughter* or son* or intergenerational* or transgenerational* or multigenerational* or generation* or caregiver* or mother-child).mp. [family setting] | 1293515 |
| 4 | ((intergeneration* or transgeneration* or multigeneration* or crossgeneration* or generation* or secondary or secondarily or tertiary or historical or vicarious or empathetic or legacy or inherited or maternal or paternal or parent* or mother* or father*) and (trauma* or PTSD or posttrauma* or psychotrauma* or abus* or maltreat* or mistreat* or adverse childhood or ACE or ACEs or childhood adversity)).ti,id. | 14171 |
| 5 | ((intergeneration* or transgeneration* or multigeneration* or crossgeneration* or generation* or secondary or secondarily or tertiary or historical or vicarious or empathetic or legacy or inherited or maternal or paternal or parent* or mother* or father*) adj4 (trauma* or PTSD or posttrauma* or psychotrauma* or abus* or maltreat* or mistreat* or adverse childhood or ACE or ACEs or childhood adversity)).mp. | 17746 |
| 6 | transgenerational patterns/ | 3515 |
| 7 | trauma/ or emotional trauma/ or posttraumatic growth/ or posttraumatic stress/ or traumatic loss/ or complex ptsd/ or exp posttraumatic stress disorder/ or exp "stress and trauma related disorders"/ | 65505 |
| 8 | 6 and 7 | 603 |
| 9 | (1 or 2) and 3 and (4 or 5 or 8) | 4676 |

Date of search: January 21 2021.

**eTable 5**. Final search strategy to identify epidemiological studies assessing the intergenerational transmission of trauma at the family- and parent-level in the Ovid PsycINFO® 1946 to Present database.

| Search term | | Yield |
| --- | --- | --- |
| 1 | (MAINSUBJECT.EXACT("Intergenerational Effects") OR NOFT(intergeneration* or transgeneration* or multigeneration* or crossgeneration* or generation* or secondary or secondarily or tertiary)) AND (NOFT(paternal* or maternal* or parent* or grandparent* or grandmother* or grandfather* or famil* or mother* or father* or child* or grandchild* or granddaughter* or grandson* or daughter* or son* or intergenerational* or transgenerational* or multigenerational* or generation* or caregiver* or "mother-child") OR MAINSUBJECT.EXACT.EXPLODE("Family Members")) | 2579 |

Date of search: January 22 2021.

**eTable 6**. Final search strategy to identify epidemiological studies assessing the intergenerational transmission of trauma at the family- and parent-level in PTSDpubs database.

| Search term | | Yield |
|---|---|---|
| 1 | ( TITLE-ABS-KEY ( intergeneration* OR transgeneration* OR multigeneration* OR crossgeneration* OR generation* OR secondary OR secondarily OR tertiary ) ) AND ( TITLE-ABS-KEY ( paternal* OR maternal* OR parent* OR grandparent* OR grandmother* OR grandfather* OR famil* OR mother* OR father* OR child* OR grandchild* OR granddaughter* OR grandson* OR daughter* OR son* OR intergenerational* OR transgenerational* OR multigenerational* OR generation* OR caregiver* OR mother-child ) ) AND ( ( TITLE ( ( intergeneration* OR transgeneration* OR multigeneration* OR crossgeneration* OR generation* OR secondary OR secondarily OR tertiary OR historical OR vicarious OR empathetic OR legacy OR inherited OR maternal OR paternal OR parent* OR mother* OR father* ) AND ( trauma* OR ptsd OR posttrauma* OR psychotrauma* OR abus* OR maltreat* OR mistreat* OR "adverse childhood" OR ace OR aces OR "childhood adversity" ) ) ) OR ( TITLE-ABS-KEY ( ( intergeneration* OR transgeneration* OR multigeneration* OR crossgeneration* OR generation* OR secondary OR secondarily OR tertiary OR historical OR vicarious OR empathetic OR legacy OR inherited OR maternal OR paternal OR parent* OR mother* OR father* ) W/4 ( trauma* OR ptsd OR posttrauma* OR psychotrauma* OR abus* OR maltreat* OR mistreat* OR "adverse childhood" OR ace OR aces OR "childhood adversity" ) ) ) ) AND ( LIMIT-TO ( DOCTYPE , "cp" ) ) | 178 |

Date of search: March 10 2021.

**eTable 7**. Final search strategy to identify epidemiological studies assessing the intergenerational transmission of trauma at the family- and parent-level in the Scopus database.

| Search term | | Yield |
|---|---|---|
| 1 | (ti((intergeneration* OR transgeneration* OR multigeneration* OR crossgeneration* OR generation* OR secondary OR secondarily OR tertiary OR historical OR vicarious OR empathetic OR legacy OR inherited OR maternal OR paternal OR parent* OR mother* OR father*) AND (trauma* OR PTSD OR posttrauma* OR psychotrauma*)) OR ab(((intergeneration* OR transgeneration* OR multigeneration* OR crossgeneration* OR generation* OR secondary OR secondarily OR tertiary OR historical OR vicarious OR empathetic OR legacy OR inherited OR maternal OR paternal OR parent* OR mother* OR father*) NEAR/2 (trauma* OR PTSD OR posttrauma* OR psychotrauma*)))) AND (ti(paternal* or maternal* or parent* or grandparent* or grandmother* or grandfather* or famil* or mother* or father* or child* or grandchild* or granddaughter* or grandson* or daughter* or son* or intergenerational* or transgenerational* or multigenerational* or generation* or caregiver* or mother-child) or ab(paternal* or maternal* or parent* or grandparent* or grandmother* or grandfather* or famil* or mother* or father* or child* or grandchild* or granddaughter* or grandson* or daughter* or son* or intergenerational* or transgenerational* or multigenerational* or generation* or caregiver* or mother-child)) AND noft(intergeneration* or transgeneration* or multigeneration* or crossgeneration* or generation* or secondary or secondarily or tertiary) | 1475 |
| 2 | Limit to doctoral dissertations | 759 |

Date of search: March 10 2021.

**eTable 8**. Final search strategy to identify epidemiological studies assessing the intergenerational transmission of trauma at the family- and parent-level in the ProQuest Dissertations and Theses database.

| Category | Extracted data |
|---|---|
| Basic study information | 1. Study design (cross-sectional, retrospective cohort, prospective cohort, or interventional)<br>2. Geographic location<br>3. Study setting (clinic, community, other)<br>4. Funding source<br>5. Sample size at baseline<br> 1. Gender distribution<br> 2. Mean and standard deviation of age (if unreported, age range)<br>6. Theoretical framework of intergenerational trauma (for example: attachment theory, family systems theory, family functioning theory, etc.).<br>7. Study definition of intergenerational transmission of trauma |
| Population characteristics | 8. Generations observed (G1, G2, G3) and relationships observed (G1 to G2; G1 to G3; G2 to G3)<br>9. Information on G1 traumatic event:<br> 1. Reported definition of G1 trauma<br> 2. Discrete or continual event (if continual, information on frequency and time range)<br> 3. Age of occurrence<br> 4. Measurement tool used to measure traumatic event |
| Exposure, mediator, moderator, and outcome information | 10. Mediating factors, for each eligible factor reported:<br> 1. Level of mediation (whether G1 to G2; G1 to G3; G2 to G3 transmission)<br> 2. Description of factor<br> 3. What path is being mediated<br> 4. Measurement tool(s)<br> 5. Timing of measurement(s) in relation to child's age<br> 6. Proportion of sample that experienced mediator versus comparator<br> 7. Impact on G2 or G3 psychological symptoms/functioning:<br> 1. Description of outcome(s)<br> 2. Measurement tool(s) and results<br> 3. Timing of measurement(s) in relation to child's age<br> 8. If mediation analyses performed:<br> 1. Sample size in analysis, analysis method (example: Baron and Kenny, structural equation modeling, etc.), standardized regression coefficient(s) for direct and indirect effects, significance test (example: Sobel), respective statistical significance level(s), covariates adjusted for, correlation matrix.<br>11. Moderating factors, for each eligible factor reported:<br> 9. Level of mediation (whether G1 to G2; G1 to G3; G2 to G3 transmission)<br> 10. Description of factor<br> 11. What path is being moderated<br> 12. Measurement tool(s)<br> 13. Timing of measurement(s) in relation to child's age<br> 14. Proportion of sample that experienced moderator/breakdown of levels<br> 15. Impact on F1 or F2 psychological symptoms/functioning:<br> 1. Description of outcome(s)<br> 2. Measurement tool(s) and results<br> 3. Timing of measurement(s) in relation to child's age<br> 16. If statistical model performed: sample size in analysis, analytic method, effect estimate(s) and respective statistical significance level(s), adjusted covariates |

**eTable 9**. Draft data extraction items.

| Studies and reference(s) | Study design | Population and study setting | N at baseline | Age and gender distribution | Concept of G1 trauma (measure) | Mediating factors (measure, timepoints, effect estimates and significance levels; covariate adjustment) | Moderating factors (measure, timepoints, effect estimates and significance levels; covariate adjustment) | G2 mental health outcomes (measure, timepoints) |
|---|---|---|---|---|---|---|---|---|
| *Intergenerational transmission from G1 to G2* | | | | | | | | |
| Study A… | | | | | | | | |
| Study B… | | | | | | | | |
| *Intergenerational transmission from G2 to G3* | | | | | | | | |
| Study C… | | | | | | | | |
| Study D… | | | | | | | | |
| *Intergenerational transmission from G1 to G3* | | | | | | | | |
| Study E… | | | | | | | | |

**eTable 10**. Draft table skeleton to provide overview of included studies and their respective results on mediators and/or moderators, sub-divided by generational transmission type.

| Category | Psychosocial factor | Definitions, measurement tools reported and in which studies |
|---|---|---|
| *Intergenerational transmission from G1 to G2* | | |
| G2 individual characteristics | Perceived transmission of trauma burden | |
| G1 individual characteristics | Severity of parental symptomatology and mental distress | |
| G1 relational characteristics | Parenting and attachment quality | |
| | Maladaptive parenting styles | |
| | Harsh parenting | |
| | Diminished parental emotional availability | |
| | Child maltreatment | |
| Family-level factors | Family structure | |
| | Decreased family functioning | |
| | Accumulation of family stressors | |
| | Dysfunctional intra-family communication styles | |
| | Environmental factors | |
| *Intergenerational transmission from G2 to G3* | | |
| …. | | |
| *Intergenerational transmission from G1 to G3* | | |
| …. | | |

**eTable 11**. Draft table skeleton to present consolidated mediation constructs. Examples were inspired from factors in recent scoping reviews in anticipation of findings.(1, 2, 5, 15, 21)
